# Smartphone-Based Symbol-Digit Modalities Test Reliably Measures Cognitive Function in Multiple Sclerosis Patients

**DOI:** 10.1101/2020.03.09.20033316

**Authors:** Linh Pham, Thomas Harris, Mihael Varosanec, Peter Kosa, Bibiana Bielekova

## Abstract

Limited time for patient encounters prevents reliable evaluation of all neurological functions in routine clinical practice. Quantifying neurological disability in a patient-autonomous manner via smartphones may remedy this problem, if such tests provide reliable, disease-relevant information.

We developed a smartphone version of the cognitive processing speed test, the Symbol-Digit Modalities Test (SDMT), and assessed its clinical utility. The traditional SDMT uses identical symbol-number codes, allowing memorization after repeated trials. In the phone app, the symbol-number codes are randomly generated.

In 154 multiple sclerosis (MS) patients and 39 healthy volunteers (HV), traditional and smartphone SDMT have good agreement (Lin’s coefficient of concordance [CCC] = 0.84) and comparable test-retest variance. In subjects with available volumetric MRI and digitalized neurological examinations (112 MS, 12 HV), the SDMT scores were highly associated with T2 lesion load and brain parenchymal fraction, when controlled for relevant clinical characteristics. The smartphone SDMT association with clinical/imaging features was stronger (R^2^ = 0.75, p < 0.0001) than traditional SDMT (R^2^ = 0.65, p < 0.0001). In the longitudinal subcohort, improvements from testing repetition (learning effects), were identifiable using non-linear regression in 14/16 subjects and, on average, peaked after 8 trials. Averaging several post-learning SDMT results significantly lowers the threshold for detecting true decline in test performance.

In conclusion, smartphone, self-administered SDMT is a reliable substitute of the traditional SDMT for measuring processing speed in MS patients. Granular measurements at home increase sensitivity to detect true performance decline in comparison to sporadic assessments in the clinic.

## Introduction

With an increasingly aging population, the prevalence of chronic neurological diseases is projected to increase dramatically. Most countries already experience a shortage of neurologists. In the United States, the average demand for neurologists is expected to be greater than supply by 20% or more by s2025 (1).

Current solutions of shifting diagnosis and care for neurological patients to primary care providers, while simultaneously decreasing the length of individual patient-encounters for neurologists, lead to delay or mistakes in diagnoses and suboptimal patient outcomes. Indeed, a comprehensive neurological examination takes 40-60 minutes to perform and years to master. Consequently, such a demanding exam is rarely performed in contemporary clinical practice, depriving patients with chronic neurological diseases of reliable measurements of their progression slopes that would guide optimal therapeutic decisions.

An alternative solution may be to develop a surrogate of neurological examination which is accessible, reliable, and sensitive to changes in disease course. To meet this need, we created a collection of smartphone-based tests, the Multiple Sclerosis (MS) Test Suite, which contains several cognitive and motoric tests that can be administered in a patient-autonomous manner (2, 3) and are expected to recreate the most important domains of a traditional neurological examination. This would allow patients to do their testing from home and have the results streamed to their clinicians. Provided that each test in the MS Test Suite validates its reproducibility and clinical relevance against the gold standard of comprehensive neurological examination, its use may speed up the identification and referrals of neurological patients from primary care providers to neurologists, help neurologists to focus their examination on affected neurological domains, and reliably track neurological disability during the disease course.

In previous papers, we have optimized and validated tests from the MS Test Suite that focused on measuring motoric and cerebellar dysfunctions in MS patients (2, 3). In this paper, we will explore the reliability and validity of another test within the app, the Symbol-Digit Modalities Test (SDMT). The SDMT is a cognitive processing speed test traditionally administered in the clinic (4). Participants are given a key of nine symbol-digit pairs along with a sequence of symbols. They are then asked to use the key and match the symbols in the sequence to their corresponding numbers. Participants are allowed 90 seconds to complete as many matches as possible and can complete the test in the written or verbal format. In the written format, participants complete the matching by writing in the number associated with each symbol in the sequence. In the verbal format, participants say the number associated with each symbol in the sequence and test administrators write in participants’ responses. Because there is only one official form of the written SDMT, repeated administration of the test may lead to memorization of the matching code, leading to one form of the practice effect, a phenomenon that occurs when individuals’ performance improves due to repeated testing. In addition to memorization of the code, traditional written SDMT also allows participants to memorize single number-symbol relationship, search for identical symbol in the sequence and fill out corresponding number for all identical symbols, irrespective of their location.

To minimize these possibilities, which collectively introduce “noise” in the measurement of processing speed, we introduced several adaptations of the smartphone SDMT app. The symbol-digit key pairing in the smartphone app changes randomly with each trial. While the entire symbol-digit key is always displayed on the top of the screen, participants are shown the symbol that they need to match one at a time, preventing them from inputting the numbers for the identical symbol out of sequence. To input their number matching responses, participants touch the corresponding number on a number keypad (numpad). A verbal version of the smartphone SDMT uses voice recognition technologies for patients whose motoric disability prevents them from using the number keypad. Because none of our patients fulfilled this criterion, we focused on comparing numpad SDMT to the traditional, written SDMT.

In addition to these issues, there are other learning mechanisms that the smartphone app could not address: test improvement over time can occur due to lessening of the anxiety associated with performing a new test or enhancing eye-hand coordination. Biological substrate of such learning is usually formation of new or strengthening of existing synaptic circuits and occurs over several test repetitions. Consequently, this paper also formally addressed the identification of learning effect in granular longitudinal sampling, as well as test-retest variance and thresholds for identification of true decline in test performance. We evaluated these thresholds for episodic assessment of SDMT in the clinic using either traditional written SDMT or smartphone SDMT and compared them to the 4-point decline in SDMT performance that was suggested as clinically meaningful change in previous studies (5-7).

Finally, we compared the performance of the smartphone (and traditional written) SDMT to an alternative cognitive test, the Paced Auditory Serial Addition Test (PASAT-3). PASAT-3 is used in the MS functional composite (MSFC) score and is frequently administered in Phase 3 clinical trials of MS disease-modifying therapies (DMTs). In the PASAT-3, participants are told a sequence of numbers. The numbers are given at intervals of 3 seconds and participants are asked to add the most recently stated number in the sequence to the number that they heard just prior.

## Materials and methods

### App development

The smartphone SDMT was written in Java and later rewritten in Kotlin, using the latest Android Studio integrated development environment. The test is delivered as a part of an Android package (APK), can be configured remotely by “prescriptions” defined and prescribed to individual subjects by the test administrator (i.e., clinician or his/her designee), and the results are stored in an online database (Google Firebase). The use of “prescriptions” allows tailoring of the Test Suite to the disability level of the test participant, which is advantageous in longitudinal applications of the Tests to limit participant’s time and frustrations with performing tests that exceed the individuals’ disability levels. The smartphone SDMT uses the latest Android operating system (Android 9 and up) and the graphics are optimized for Google Pixel XL and Google Pixel 2 XL, in colorblind-friendly visualizations. The symbols are a set of Unicode characters, with the symbol-digit key and symbol sequence shuffled randomly for every test.

### Participants

This study was approved by the Central Institutional Review Board of the National Institutes of Health (NIH) and all participants signed informed consent. Healthy volunteers (HV) and MS participants enrolled in one of two protocols: Targeting Residual Activity by Precision, Biomarker-Guided Combination Therapies of Multiple Sclerosis (TRAP-MS; clinicaltrials.gov identifier NCT03109288) or Comprehensive Multimodal Analysis of Neuroimmunological Diseases in the Central Nervous System (NCT00794352). Two HV cohorts were recruited via NCT00794352: 1) HV that undergo all study procedures (including full physical and neurological examination, PASAT-3, traditional written SDMT, MRI of the brain) and serve as controls to all studies administered under this protocol, and 2) Smartphone app self-declared HV group recruited with the intent to collect longitudinal data from the larger cohort of HV that use the smartphone MS Test Suite remotely to mimic real-world situation. These HV participants signed digital informed consent that tested their comprehension of the testing instructions and associated risks/benefits directly via smartphone. Participant demographics are summarized in Table 1 and 2.

**Table 1:** Demographics table with baseline information for cross-sectional data.

| **A. Demographics** | **All Cross-Sectional Smartphone Participants (N = 193)** |  |  |  |
| --- | --- | --- | --- | --- |
|  | **PPMS (n = 54)** | **SPMS (n = 42)** | **RRMS (n = 58)** | **HV (n = 39)** |
| ***Age (year)*** |  |  |  |  |
| Mean ± SD | 58.7 ± 11.1 | 58.3 ± 10.3 | 47.3 ± 9.6 | 41.1 ± 14.0 |
| Range | 18.5 - 72.8 | 33.0 - 75.5 | 30.2 - 77.2 | 21.5 - 67.9 |
| ***Gender (% of cohort)*** |  |  |  |  |
| Female | 56 | 57 | 60 | 59 |
| Male | 44 | 43 | 40 | 41 |
| **B. Demographics** | **Cross-Sectional Smartphone Participants with Clinical Data (N = 112)** |  |  |  |
|  | **PPMS (n = 46)** | **SPMS (n = 26)** | **RRMS (n = 40)** | **HV (n = 12)** |
| ***Age (year)*** |  |  |  |  |
| Mean ± SD | 58.5 ± 10.8 | 56.7 ± 10.8 | 45.6 ± 8.3 | 48.9 ± 6.8 |
| Range | 18.5 - 72.8 | 33.0 - 75.5 | 30.2 - 66.4 | 40.1 - 60.8 |
| ***Gender (% of cohort)*** |  |  |  |  |
| Female | 61 | 58 | 62 | 75 |
| Male | 39 | 42 | 38 | 25 |
| ***NeurEx*** |  |  |  |  |
| Mean ± SD | 162.0 ± 61.0 | 164.0 ± 60.1 | 68.5 ± 48.3 | 18.4 ± 15.1 |
| Range | 34.2 - 312.0 | 67.1 - 334.0 | 0.0 - 216.0 | 0 - 54.2 |
| ***EDSS*** |  |  |  |  |
| Mean ± SD | 5.6 ± 1.0 | 5.7 ± 1.0 | 3.6 ± 1.5 | 1.9 ± 0.8 |
| Range | 3.0 - 7.0 | 4.0 - 6.5 | 0.0 - 6.5 | 0.0 - 3.0 |

**Table 2.** Demographic table with information for individuals used in written SDMT change threshold determination and app SDMT longitudinal analyses. Demographics in table A come from the second visit of the 6-months span. Demographics in table B come from the first visit (baseline) values.

| A. Demographics | Participants Included in Written Threshold Analysis (N = 112) |  |  |
| --- | --- | --- | --- |
|  | PPMS (n = 47) | SPMS (n = 33) | RRMS (n = 32) |
| **Age (year)** |  |  |  |
| Mean ± SD | 57.3 ± 11.5 | 57.0 ± 8.2 | 47.2 ± 10.8 |
| Range | 19.0 - 70.8 | 38.1 - 71.2 | 29.8 - 77.7 |
| **Gender (% of cohort)** |  |  |  |
| Female | 60 | 67 | 62 |
| Male | 40 | 33 | 38 |
| **NeurEx** |  |  |  |
| Mean ± SD | 159.0 ± 69.9 | 178.0 ± 48.2 | 64.8 ± 44.5 |
| Range | 54.3 - 328.0 | 74.4 - 288.0 | 4.2 - 165.0 |
| **EDSS** |  |  |  |
| Mean ± SD | 5.7 ± 1.1 | 6.1 ± 0.8 | 3.5 ± 1.5 |
| Range | 2.5 - 7.5 | 4.0 - 7.5 | 1.0 - 6.5 |

| B. Demographics | Longitudinal Smartphone Participants (N = 16) |  |  |  |
| --- | --- | --- | --- | --- |
|  | PPMS (n = 8) | SPMS (n = 5) | RRMS (n = 2) | HV (n = 1) |
| **Age (year)** |  |  |  |  |
| Mean ± SD | 59.0 ± 12.9 | 59.5 ± 4.9 | 54.1 ± 1.3 | 63.1 |
| Range | 28.2 - 69.3 | 54.2 - 64.9 | 53.2 - 55.0 | na |
| **Gender (% of cohort)** |  |  |  |  |
| Female | 38 | 40 | 0 | 100 |
| Male | 62 | 60 | 100 | 0 |
| **NeurEx** |  |  |  |  |
| Mean ± SD | 197.0 ± 112.0 | 164.0 ± 103.0 | 46.1 ± 18.2 | na |
| Range | 88.9 - 436.0 | 67.1 - 334.0 | 33.2 - 59.0 | na |
| **EDSS** |  |  |  |  |
| Mean ± SD | 6.1 ± 1.2 | 5.4 ± 1.0 | 3.0 ± 0.7 | na |
| Range | 4.5 - 8.0 | 4.0 - 6.5 | 2.5 - 3.5 | na |

### Tests administration and data collection

In cross-sectional testing, the traditional written SDMT, PASAT-3, and smartphone SDMT were administered to MS patients and HV during the same visit day (test sitting). Participants were instructed by a trained administrator on how to perform each test and completed a practice trial prior to testing. On the smartphone SDMT, if a test trial had technical errors (e.g., test interruption), participants had the option to retake the trial. The smartphone SDMT required participants to use a number keypad to input their number matching response to a given symbol (Fig. 1). To ensure that the smartphone and written SDMT results were comparable, participants used their dominant hand for inputting the answer on the smartphone test. Initially, each test sitting required participants to complete two trials of the smartphone SDMT. This was eventually reduced to one trial in the interest of time.

**Figure 1.**
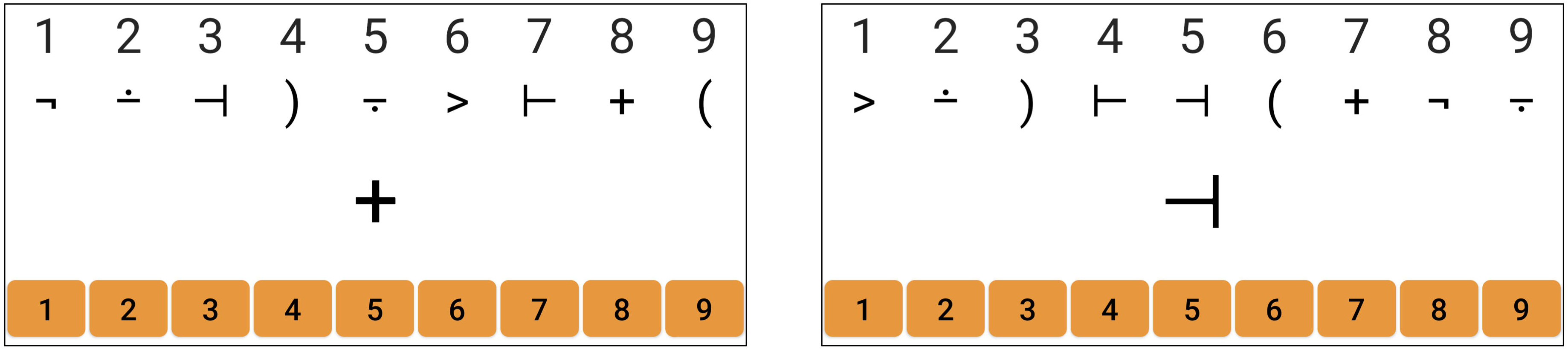
The interface for the app based SDMT. The symbol-digit key changes with every trial. During the test, participants are only seeing one symbol at a time and use a number keypad (orange buttons) to input the matching number to the symbol being shown.

MS patients who expressed interests were provided with a smartphone pre-loaded with MS Test Suite. Patients were then instructed on how to navigate the app prior to home testing. They were asked to test, at minimum, once per week and at around the same time of day (morning, night, etc.). MS and HV participants who completed at least 20 sittings comprised the longitudinal testing cohort. Cross-sectional participants (Table 1B: 112 MS; 12 HV) who underwent neurological exams by MS-trained clinicians had their disabilities documented in NeurEx, an iPad-based app that accurately measures disabilities in 17 different neurological domains (8). A disability score is derived for each domain, with higher score indicating more disability in that domain.

### Volumetric brain MRI analyses

Brain MRIs were performed on 3T Signa units (General Electric, Milwaukee, WI) or 3T Skyra (Siemens, Malvern, PA). To calculate T2 Lesion Load (T2LL) as well as Brain Parenchymal Fraction (BPFr), T1 magnetization-prepared rapid gradient-echo (MPRAGE) or fast spoiled gradient-echo (FSPGR) and T2 weighted three-dimensional fluid attenuation inversion recovery (3D FLAIR) were acquired. Detailed settings on the 3TA Signa scanner for SPGR sequences, were run with two flip angles (Repetition Time (TR) 7.8 ms; Echo Time (TE) 3 ms; Flip Angle (FA) 3° and 17°; 1-mm isotropic resolution; acquisition time (TA) 4 minutes). On 3TD, Siemens, the MRI scan included 3D-GRE with two flip angles for quantitative T1 (qT1) mapping (TR 7.8 ms; TE 3 ms; FA 3° and 18°; 1mm isotropic resolution, TA 3.5 minutes. These sequences were used to generate volumetric data.

Raw unprocessed but locally anonymized and encrypted T1 - MPRAGE or T1 - FSPGR and T2 - 3D FLAIR DICOM files, as inputs, were analyzed by an automated segmentation algorithm called LesionTOADS (9) which is implemented into a cloud based service for medical image processing, QMENTA (www.qmenta.com). In detail, the uploaded sequences are anterior commissure-posterior commissure (ACPC) aligned, rigidly registered to each other and skull stripped (the T1 image is additionally bias-field corrected). The segmentation is performed by using an atlas-based technique combining a topological and statistical atlas resulting in computed volumes for each segmented tissue in mm^3^.

### Data blinding protocol

All patients received a random code that allows their results to be blinded for the relevant parties. Smartphone app, written SDMT, and PASAT-3 were performed by technicians who were blinded to neurological examination results. Clinicians performing neurological examinations while being blinded to MRI and smartphone results. MRI analyses were performed by investigators who were blinded to clinical examination and smartphone outcomes.

### Implementation of non-linear regression

Non-linear regression was used for two purposes: 1) To identify the minimum amount of time needed for smartphone SDMT to achieve comparable performance to traditional written SDMT and 2) To identify practice effect (if there is any) in the longitudinal tester subgroup. An explanation of non-linear regression can be found in Figure 2A.

**Figure 2.**
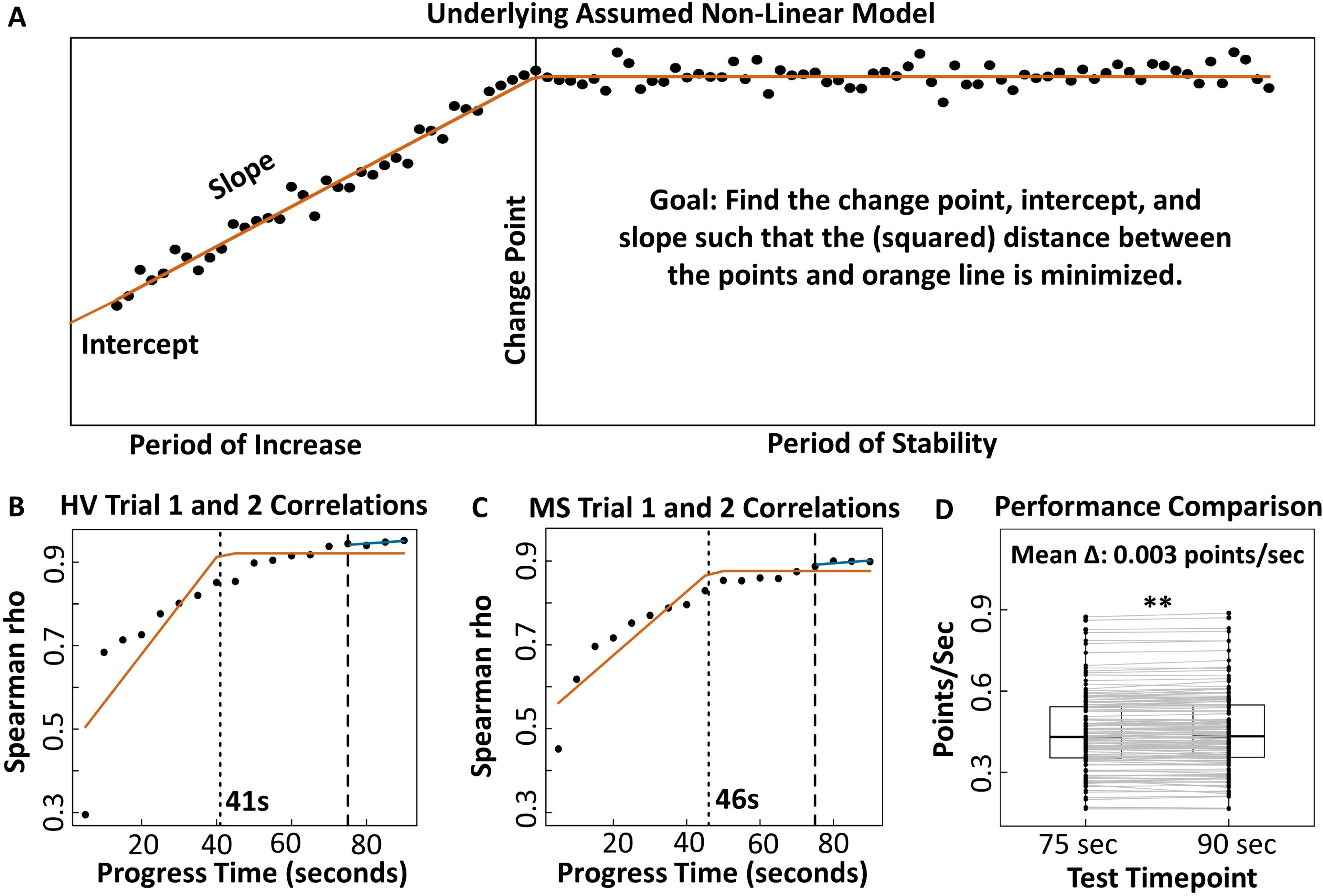
Minimum time needed for reliable app based SDMT performance. **(A)** Demonstration of how the non-linear regression model works to find the inflection/change points. **(B-C)** Plots of correlation in the scores between two trials (in the same test sitting) as the testing time progresses for HV and MS. Orange lines represent the underlying non-linear regression models fitted to the data. Dotted lines indicate the minimum time needed for reliable performance based on the non-linear regression algorithm. Dashed lines point to the correlation coefficients at 75 seconds, or the time constraint that was eventually implemented. Blue lines indicate that after 75 seconds, the correlation continues to slightly improve. **(D)** Comparison of testing speed, in points per seconds, within the same trial at 75 and 90 seconds. Individuals show strong evidence of increasing their speed towards the end of the test (p < 0.001) but the change of 0.003 points/sec is clinically insignificant.

### Identification of minimum testing time on the smartphone SDMT

Time is an essential determinant in the utility of a clinical test: because the goal of the MS Test Suite is to recreate the neurological examination, it consists of multiple different tests. If administration of these tests takes too long, subjects will be discouraged to complete the Suite. Because we could not find published rationale for the selection of 90 seconds for the SDMT test, we used a data-driven approach during test development to determine the shortest length of the test that still provides reliable information.

For this analysis, we used raw (no outliers removed) cross-sectional test-retest reliability data from individuals who completed two 90-seconds trials in the same test sitting. Spearman correlation coefficients of test results (calculated as number of correct answers at each test duration) were generated for every 5 seconds of the duration of the two trials. As expected, these correlations increased with the duration of the test, but eventually plateaued around Rho 0.9 in both HV and MS cohorts (Fig. 2B-C). Using non-linear regression, we identified 2 major infliction points in these time-lapse data: major infliction point at 41 seconds test duration in HV and 46 seconds in MS. However, the correlation was still improving after these times, albeit at much slower pace (Fig. 2B-C). Therefore, to maximize test-retest reliability we selected second infliction point at 75 seconds of test duration, when the test-retest correlation maximized. We then used the 75 seconds test duration for all subsequent testers.

To formally test that shortening of app based SDMT time to 75 seconds will provide comparable results to traditional, 90 sec SDMT, we analyzed the within-subject difference in the test performance (correct answers/sec) between 75 seconds and 90 seconds. A Wilcoxon signed-rank test shows strong evidence (p < 0.001) that an improvement of 0.003 points/sec occurs between the 75 seconds and 90 seconds time points within the test (Fig. 2D). This change is clinically insignificant in comparison to between participant’s variance that ranges from 0.2 to 0.9 correct answers per second.

To keep the smartphone SDMT results comparable despite the change in time limit, all smartphone SDMT scores are re-calculated using the following formula:

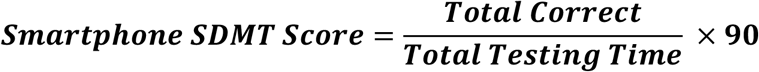

### Pre-processing of the data: Identification and adjustment of outlier values

For any test sittings that had two back-to-back trials, the trials’ score difference and mean were calculated. The results were visualized using a Bland-Altman plot (Fig. 3A). The mean difference and the limits of agreement were calculated using the protocol described by Parker et al. (10) for data with unbalanced but repeated measurements across different subjects. The equations used for calculating these values can be found in Suppl. 1. We observed that the average difference between trial 1 and 2 in all test sittings with two trials is 0.19 correct answers recalculated per 90 seconds of test (Fig. 3A). Again, this change is clinically insignificant, indicating that on average the trials 1 and 2 are comparable. However, the variance of these test-retest data greatly exceeds the 4-point difference that is currently accepted as clinically meaningful decline in SDMT performance (Fig. 3A). We get back to this observation later in the results.

**Figure 3.**
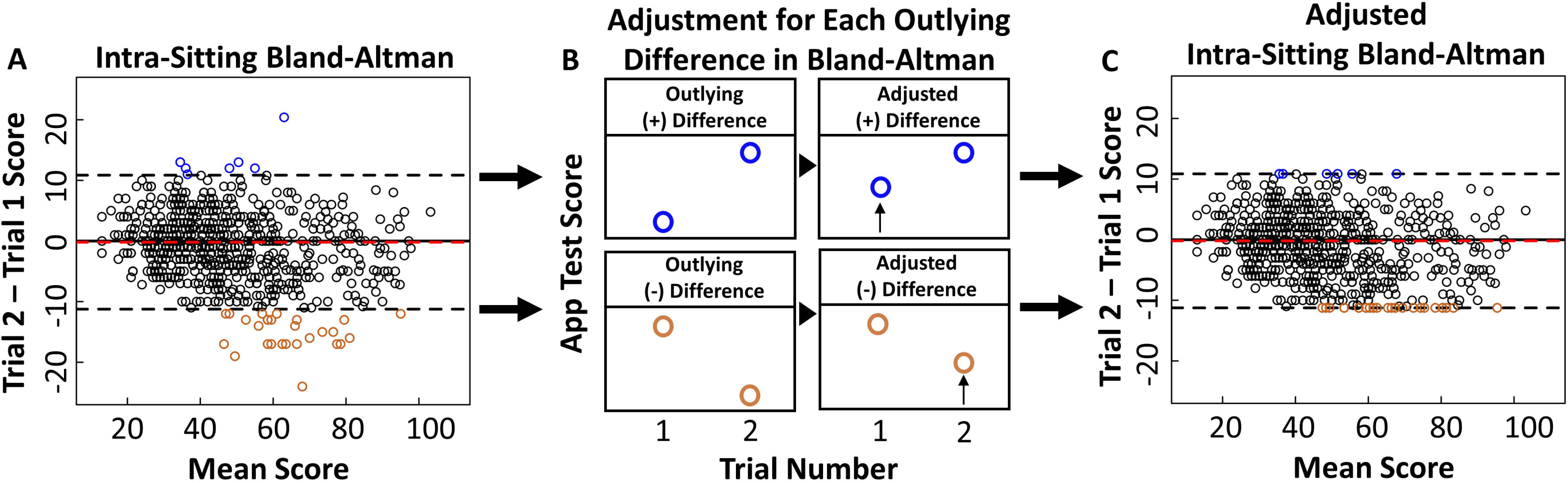
Outlier removal procedure for test sittings with two trials. **(A)** Score differences and means for test sittings with two trials are shown on a mixed-effects Bland-Altman plot. The limits of agreement are −11.24 and 10.85 and the mean difference is −0.19. Sittings with differences that fall outside of the limits of agreement, indicated by blue and orange points, are considered to have one trial that is an outlier. **(B)** In the sittings with outlier trials, the lower performing trial’s score is increased to reach the limits of agreement thresholds. **(C)** The original mixed-effects Bland-Altman plot, now shown with the outlying differences after score adjustment.

Outliers were identified as sittings where the difference fell outside of the Bland-Altman limits of agreement. Because the participants demonstrated ability to perform the test at the higher level, we arbitrarily assigned an outlier status to the lower performing score from two back-to-back trials. However, rather than completely excluding the lower-performing outlier trial, which would introduce bias in comparison to all other subjects where the SDMT value was calculated as average of two attempts, we increased the score of the outlier test till the score difference reached the closest limit of agreement on the Bland-Altman plot (Fig. 3B – C). All data pre-processing and analyses were done in the statistical software R (11). A list of all packages used in this paper can be found in the references (12-31). All analysis scripts can be found in the Supplementary Materials.

### Statistics

The mean comparison tests used throughout this paper are non-parametric Wilcoxon signed-rank test (comparing paired data) and Wilcoxon rank-sum test (comparing unpaired data). All correlation analyses are conducted using Spearman correlation coefficients and associations that control for confounding variables are conducted using elastic net regression. More information on elastic net regression can be found in Suppl. 5. A conservative significance threshold of 0.01 is used for all comparison, correlation, and association analyses.

For agreement analyses between two different methods that measure the same outcome, the Lin’s coefficient of concordance (CCC) was used (32). The CCC is calculated based on the strength of correlation between the two methods and the degree in which their regression line follows a 1:1 trajectory. The closer the CCC is to 1, the higher the agreement between the two methods.

For reliability analyses across multiple time points, such as in the case of the longitudinal smartphone SDMT data, an intraclass correlation coefficient (ICC) was employed. The ICC value determines how much the variance within the overall data is due to within-individual clustering comparing to between-individual clustering (within / [between + within]). ICC values range from 0 to 1. The closer the ICC value is to 1, the more the overall variance of the data is explained by clustering within the same individual and therefore, the higher the reliability of the data within the same test taker. The sitting number was used as the fixed effect in this model and the equation used for calculating the ICC value can be found in Suppl. 9. The ICC results using days from the first trial (time) as the dependent variable can be found in Suppl. 10-11.

### Training and validation cohorts for elastic net modeling

In the elastic net regression analyses, the cross-sectional data was divided into training and validation cohorts. The training cohort was used to obtain the regression equation that describes the relationship between the selected SDMT outcomes and their associated smartphone, clinical, and/or MRI variables. The validation cohort was used to assess how well the trained equation can explain, or generalize, the relationship in a new group of individuals. The training cohort had 2/3 of the subjects in the cross-sectional data and the validation cohort had 1/3 of the subjects. The cohorts were divided to have similar distributions in age, smartphone SDMT, and written SDMT scores (Suppl. 6).

## Results

### Cross-sectional analyses

#### Smartphone SDMT can differentiate HV from MS cognitive processing speed and replace the written SDMT

To determine if the smartphone SDMT is a valid measurement of cognitive processing speed, we assessed its ability to differentiate between HV and MS performance – a feature which is well-documented in the traditional, investigator-administered SDMT (33). While there is an overlap in the smartphone SDMT performance range for HV and MS (HV: 23.5 – 92.4 points; MS: 6.0 – 76.5), the Wilcoxon rank-sum test presents convincing evidence that the HV and MS cohorts perform differently on the smartphone SDMT (p < 0.0001, Fig. 4A). The HV cohort’s median performance is 54 points and the MS cohort’s median performance is 34.8 points.

**Figure 4.**
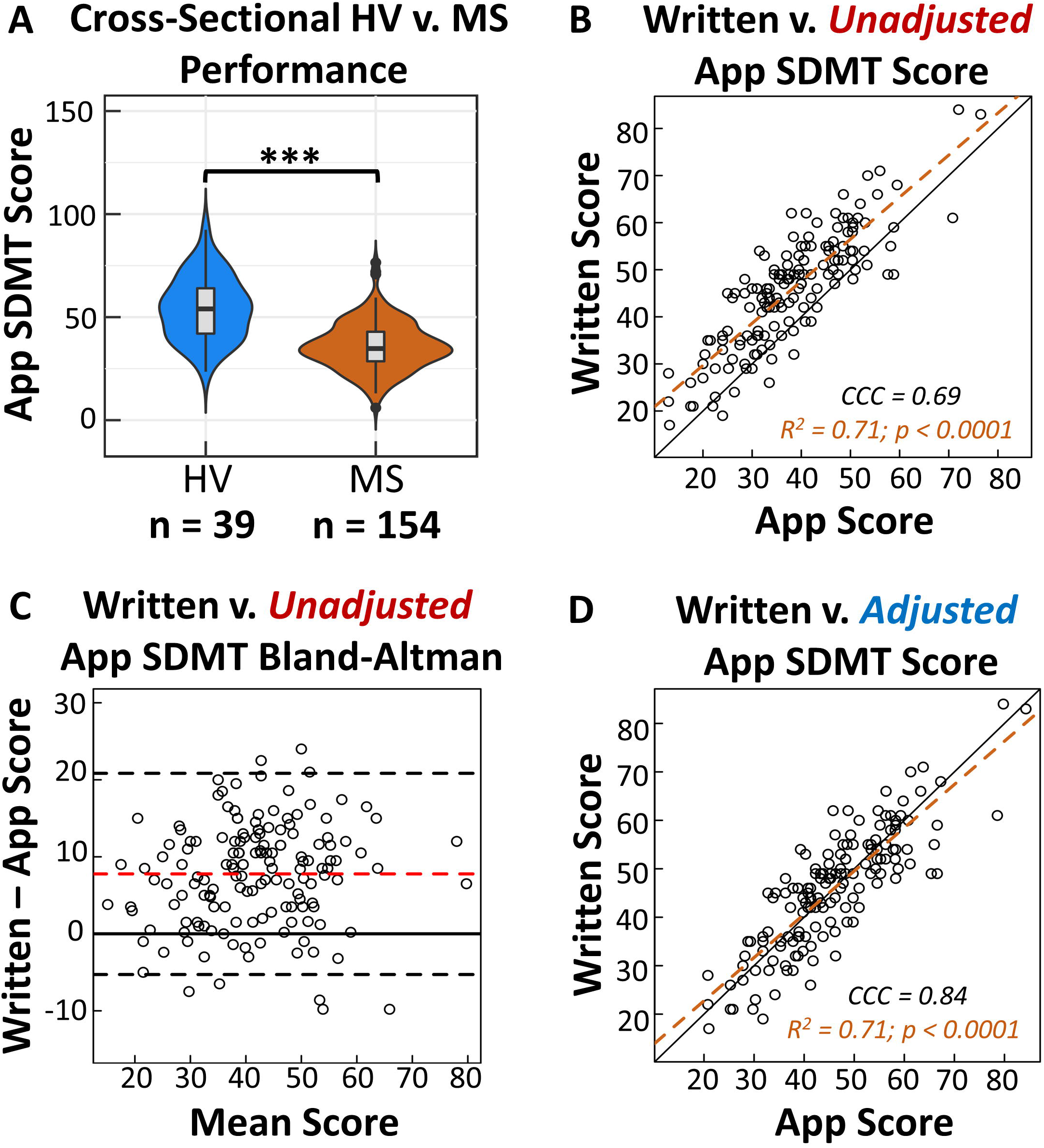
Analysis of app based SDMT validity. **(A)** HV generally perform better than MS on the app SDMT (p < 0.0001). The number of participants for each cohort are shown below the plot. **(B)** Assessment of agreement between the raw app based score and the written score. The black solid line is the 1:1 concordance line. The orange dashed line is the regression line. **(C)** Bland-Altman plot for the written and the app scores. The limits of agreement are −5.29 and 20.84. On average, individuals perform 7.8 points higher on the written test. **(D)** The plot of concordance between the written and app scores, after adjusting for the 7.8 points difference.

Next, we asked if the smartphone SDMT is directly comparable to investigator administered, written SDMT. While the smartphone and written results show strong evidence of association (R^2^ = 0.71, p < 0.0001), we observed a positive bias in the direction of the written scores, indicating that subjects were generally scoring higher on the written test (CCC = 0.69, Fig. 4B). Visualizing this bias on a Bland-Altman plot shows that on average, individuals across different levels of SDMT performance do better on the written version by 7.8 points (Fig. 4C).

There are several possible sources of this bias, first two of which we recognized as a likely drawback of written SDMT and deliberately addressed in the smartphone SDMT design: 1. Memorization of the symbol-digit code: Because vast majority of our MS patients already experienced traditional written SDMT before, it is possible that they memorized the symbol-digit code. 2. Filling digits for a specific symbol out of sequence. 3. Written test may be easier to see and perform due to larger numbers. 4. There could have been an element of anxiety in using the new test. Irrespective of these causes, to directly compare written and smartphone SDMT results, we must add 7.8 points to all smartphone scores. This adjustment increased the CCC value between the two methods to 0.84 (34) (Fig. 4D).

#### SDMT associates with brain atrophy and lesions volume when controlling for confounding variables, with smartphone SDMT achieving higher correlation in comparison to written SDMT

Although traditional SDMT is the “state of the art” test in contemporary practice, the possible drawbacks we identified and addressed in smartphone SDMT app design leaves a possibility that app SDMT may better reflect decline in reaction time caused by MS-mediated destruction of central nervous system (CNS) tissue.

To test this hypothesis, we assessed Spearman correlations between app based SDMT, traditional SDMT results, state of the art clinical measures and brain volumetric measures previously associated with cognitive functions and CNS tissue destruction in MS (Fig. 5). Disability measures were derived from NeurEx, which is a freely-available iPad app that allows clinicians to conveniently document the entire neurological examination in a very short time using intuitive touch interface (35). This digitalization of the neurological examination streamed to a secured cloud database allows for automated computation of major disability scales used in neuroimmunology, as well as provides neurological subsystems and limb-specific disability data for research applications. Brain volumetric measures were derived from brain MRIs acquired within the same clinic visits. We focused our analysis on BPFr and T2LL, which showed correlations to traditional SDMT results in previous study (36). As a comparison to written and app based SDMT, we included PASAT-3, another cognitive test broadly used in MS. Age was included in the correlation analyses as a potential covariate.

**Figure 5.**
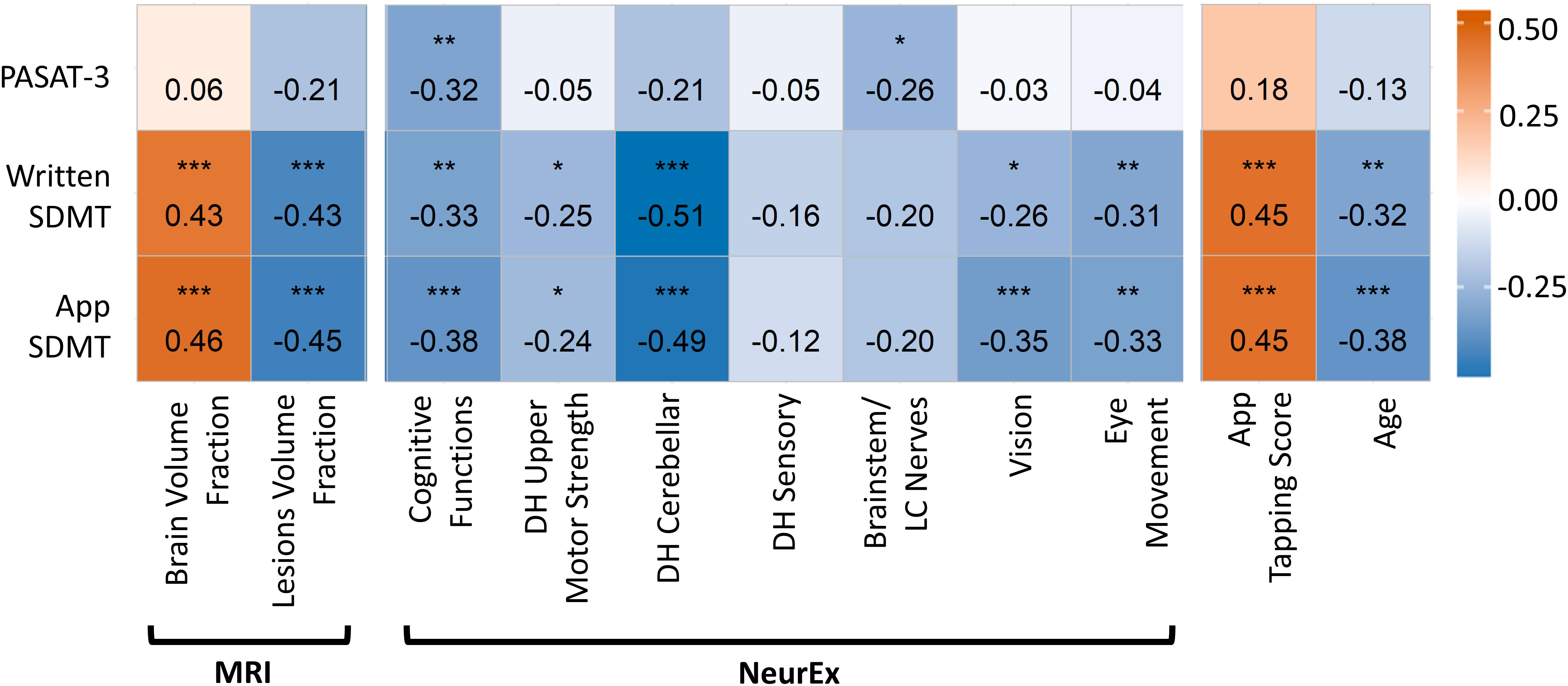
Spearman correlation coefficients between cognitive tests and outcomes from brain MRI, NeurEx features, smartphone tapping test, and age. p-values in the matrix are indicated as followed: *p < 0.01, **p < 0.001, ***p < 0.0001. Smartphone and written SDMT show evidence of correlation to the same set of features. PASAT-3 correlates with NeurEx cognitive and brainstem functions but does not correlate with volumetric MRI features.

These analyses validated that written SDMT results positively correlate with BPFr and negatively correlate with T2LL and cognitive functions subsystem of NeurEx (Fig. 5). For most of these, app based SDMT exhibited stronger correlation coefficients and lower p-values in comparison to written SDMT. In contrast, PASAT-3 did not correlate with MRI volumetric measures, even though it exhibited similar negative correlation to NeurEx cognitive functions subscore as written SDMT. Additional differences between PASAT-3 and both SDMT tests resided in the differences in correlations with motoric/cerebellar dominant hand (DH) functions and vision/eye movement subscores of NeurEx, which were significant for both SDMT tests, but non-significant for PASAT-3. In contrast PASAT-3 had mild, but statistically significant correlation with brainstem/lower cranial nerve subscores of NeurEx, which was absent for 2 SDMT tests. Thus, we conclude that observed correlations with NeurEx subdomains are in full agreement with the fact that PASAT-3 utilizes only hearing and voice (i.e., captured by brainstem/lower cranial nerves subscore of NeurEx) and higher cognitive functions, while written SDMT performance relies on vision, eye/hand coordination and manual dexterity of the DH. The stronger correlation with vision and eye movement subscores of NeurEx observed for app based SDMT versus traditional written SDMT suggests that smaller screen of smartphone SDMT in comparison to paper version provides a greater challenge to visual system.

The observed negative correlations of SDMT scores with dexterity and visual functions suggested that we may better isolate true cognitive disability aspect of SDMT results if we adjust for disability in the visual or motoric/cerebellar subsystems. To test this hypothesis, we used elastic net (EN) regression, a type of multiple linear regression that produces association coefficients while accounting for multicollinearity among the predictor variables. In these regression models, the smartphone or written SDMT performance was used as the response variable. Because BPFr and T2LL correlated stronger to the SDMT results than the measured cognitive functions subscores in NeurEx, we chose these MRI features as predictor variables reflecting CNS tissue destruction in the telencephalon as an anatomical substrate of the cognitive disability in MS. Any remaining NeurEx factors which showed strong evidence of correlation to these results were included as covariates in the models.

Because mathematical algorithms that use multiple variables to model the outcome are susceptible to overfitting (i.e., the algorithms may use “noise” represented by random associations of measured variables to the outcome that are not based on biological relationships and therefore may not be reproducible), the general validity of such models must be tested in an independent validation cohort. Therefore, we randomly divided our cohort to training (2/3) and validation (1/3) subcohort (See Suppl. 6).

In the *training cohort*, the EN models showed that BPFr and T2LL were most associated with the smartphone and written SDMT results, when controlling for the influences of other disabilities and age (Fig. 6A). While DH cerebellar function, motoric strength, and age were important covariates for both tests, the EN deemed vision and eye movement dysfunctions to be important only in the smartphone SDMT results, consistent with stronger Spearman correlations observed for app-SDMT and these visual functions.

**Figure 6.**
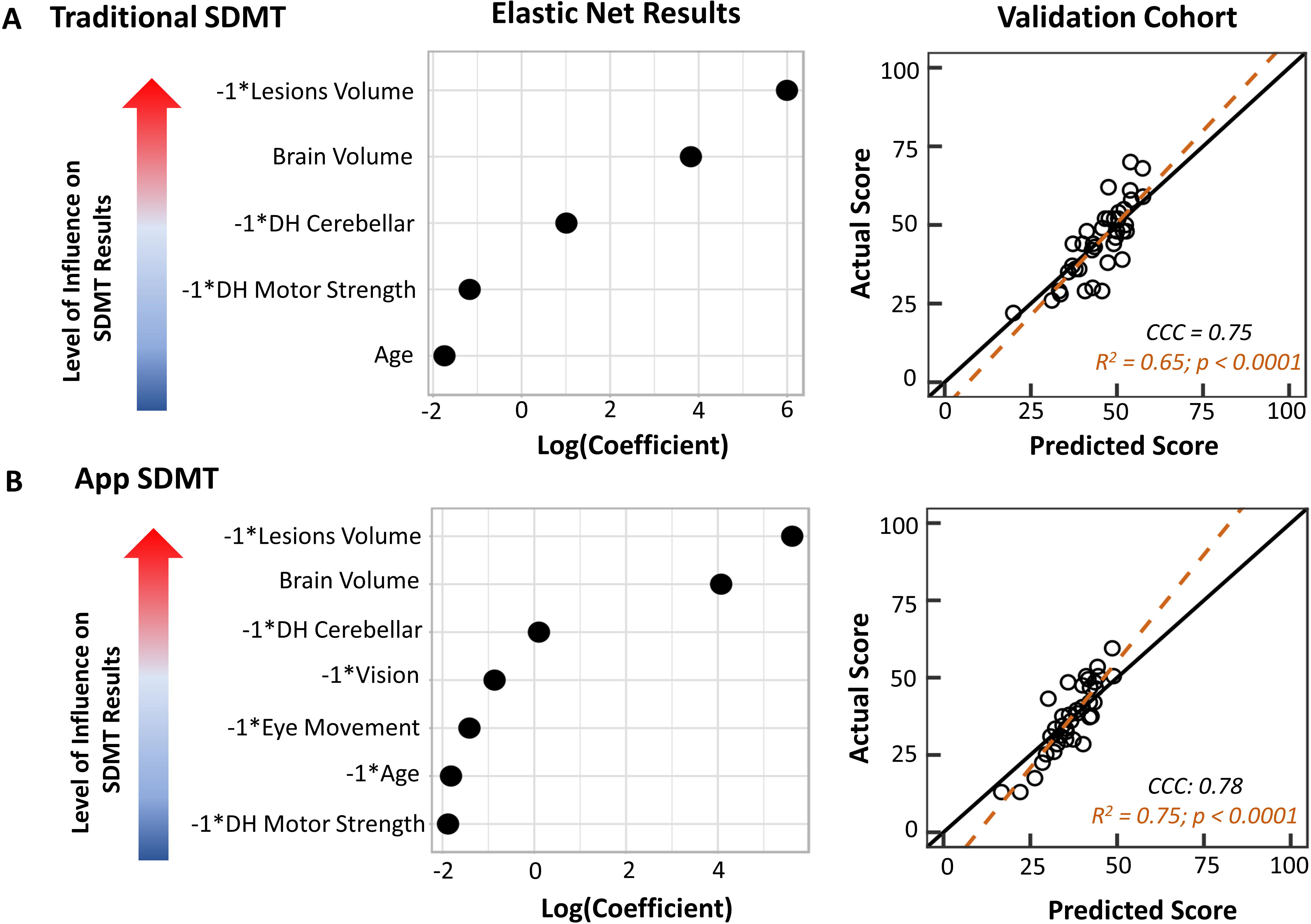
Clinical associations for traditional/written and app SDMT. **(A)** Elastic net regression shows that when controlling for relevant motoric factors and age, brain parenchymal fraction and T2 lesions load are highly associated with the written results. These factors predict a traditional SDMT score that agrees with the actual score at a CCC of 0.75. **(B)** For the app SDMT, vision and eye movement also influenced the results. Controlling for these factors, brain parenchymal fraction and T2 lesions load are most associated with the app results. This model produces a predicted app SDMT value that agrees to the actual value at a CCC of 0.78.

When we applied resulting equations to the independent *validation cohort*, they were able to predict the smartphone results better than the written SDMT results (written: R^2^ = 0.65, p < 0.0001; smartphone: R^2^ = 0.75, p < 0.0001). This result indicates that adjusting for age and disabilities in the visual system and neurological functions that influence DH dexterity indeed better isolates CNS tissue damage that underlies slowing of reaction speed in MS.

#### Influence of motoric disabilities on the smartphone SDMT results can be controlled for using smartphone tapping score

The modeling introduced in previous section relied on clinician-derived disability measures which will not be broadly available. Thus, we asked whether we can use a surrogate to the relevant clinician-derived measures from our MS Test Suite. Although we implemented and are currently optimizing tests in MS Test Suite that may be used as surrogate of clinician-derived visual functions, we currently do not have sufficient data for tests analyses and validation of the app-based visual outcomes against gold standard of clinician-derived measures. We do have readily available DH tapping results, previously validated against clinician-derived disability scores in neurological subdomains that underlie DH dexterity (3). To demonstrate this relationship, we included correlations between DH tapping scores and SDMT results in Figure 5.

Consequently, we asked whether adjusting the SDMT scores for the number of taps available in the MS Test Suite improves EN modeling of SDMT results using relevant volumetric MRI markers of brain tissue destruction in MS. When we replaced the NeurEx scores of DH cerebellar and upper motoric strength with tapping results in the smartphone SDMT EN model, the model validated only slightly worse than the model that included the NeurEx scores (R^2^ = 0.67, p < 0.0001; Fig. 7A).

**Figure 7.**
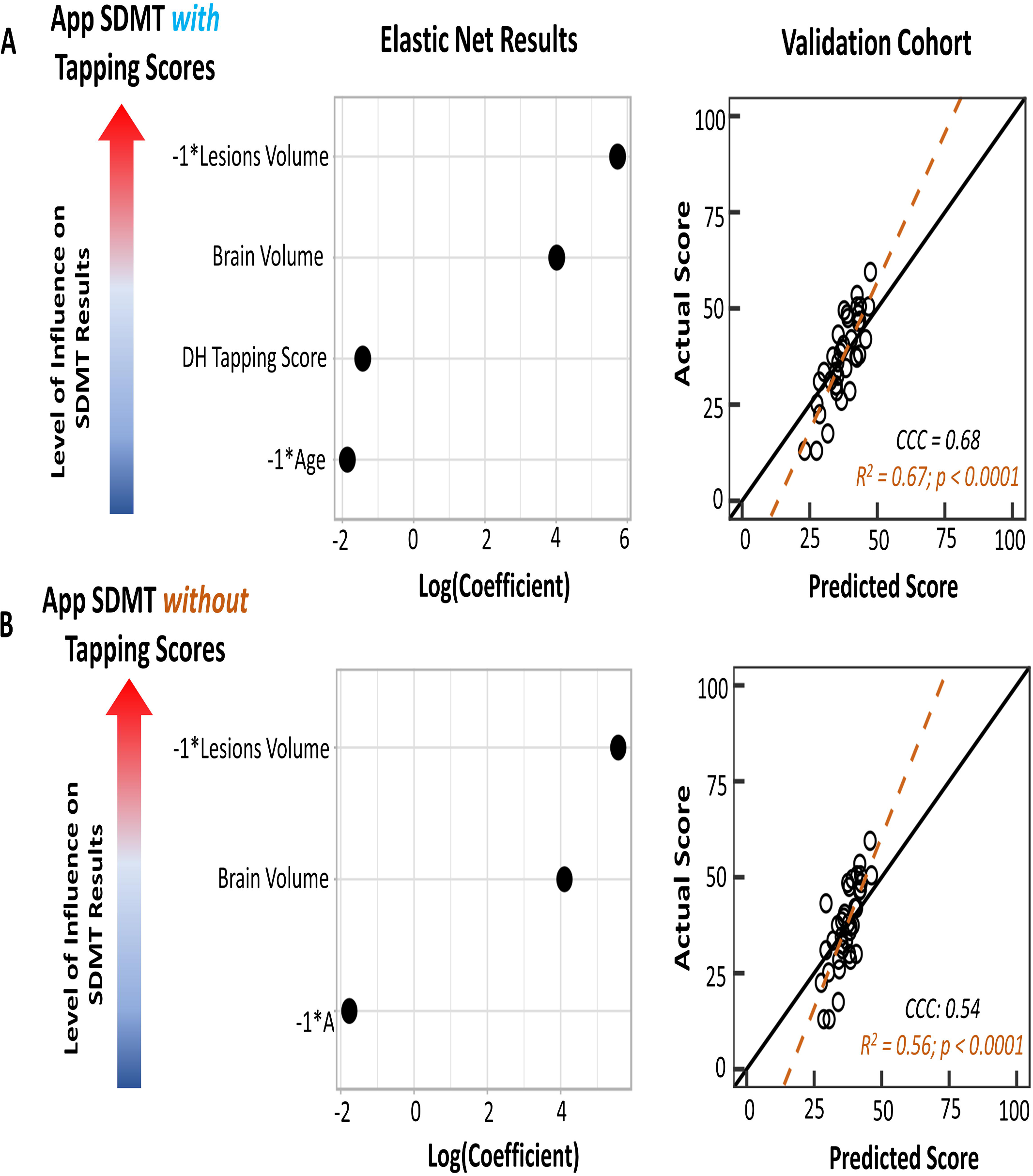
App tapping score serving as a covariate in the elastic net model. **(A)** Elastic net regression results with NeurEx cerebellar functions and upper motor strength being replaced by the app tapping score as a covariate. This model produces scores that agree with the actual scores at a CCC of 0.68. **(B)** Removing the tapping scores reduces the CCC to 0.54, showing that while tapping scores, like cerebellar and upper motor functions, have small influences on the app SDMT comparing to the MRI variables, it is important to include this covariate in the model.

Even though DH tapping score has much lower weight in EN SDMT model than the two MRI features, when we eliminated tapping score from the model (Fig. 7B) we observed a substantial decrease in model performance. We conclude that the smartphone tapping score is a good proxy for clinician-derived disability scores to control for loss of DH dexterity in evaluating the relationship between SDMT results and MS-related brain tissue destruction reflected by BPFr and T2LL.

Rewriting validated EN-based multiple linear model equation, adjusting the app based SDMT result using the following formula will produce results that explain 67% of the variance associated with MS-related structural damage to brain tissue, which is an anatomical substrate of MS-associated cognitive dysfunction.

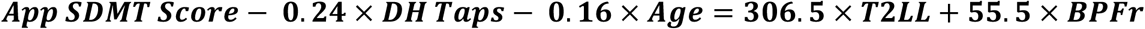

### Longitudinal analyses

#### Individual-specific practice effects are quantifiable using non-linear regression

Within our MS cohort, a subgroup of patients uses the smartphone SDMT from home on a weekly basis. This presents a unique opportunity to investigate the prevalence of practice effect in the longitudinal testing. Determining when the practice effect stops will help clinicians identify which data points from the smartphone SDMT to disregard when assessing for subsequent true decline in SDMT scores.

To begin quantifying individual practice effects on the smartphone SDMT, we first plotted the individuals’ longitudinal data to look for trends that would indicate the occurrence of practice effect (Fig. 8). This trend, which involves the smartphone SDMT performance increasing then stabilizing into a plateau that fluctuates around a mean score, was discernable when the smartphone SDMT has been completed for at least 20 test sittings. Using non-linear regression, we identified the sitting at which the performance trend switches from increasing to plateauing. This sitting is deemed to be the point where the individual’s practice effect stopped. Any trial that occurred after this point can be used to assess the individual’s true cognitive changes.

**Figure 8.**
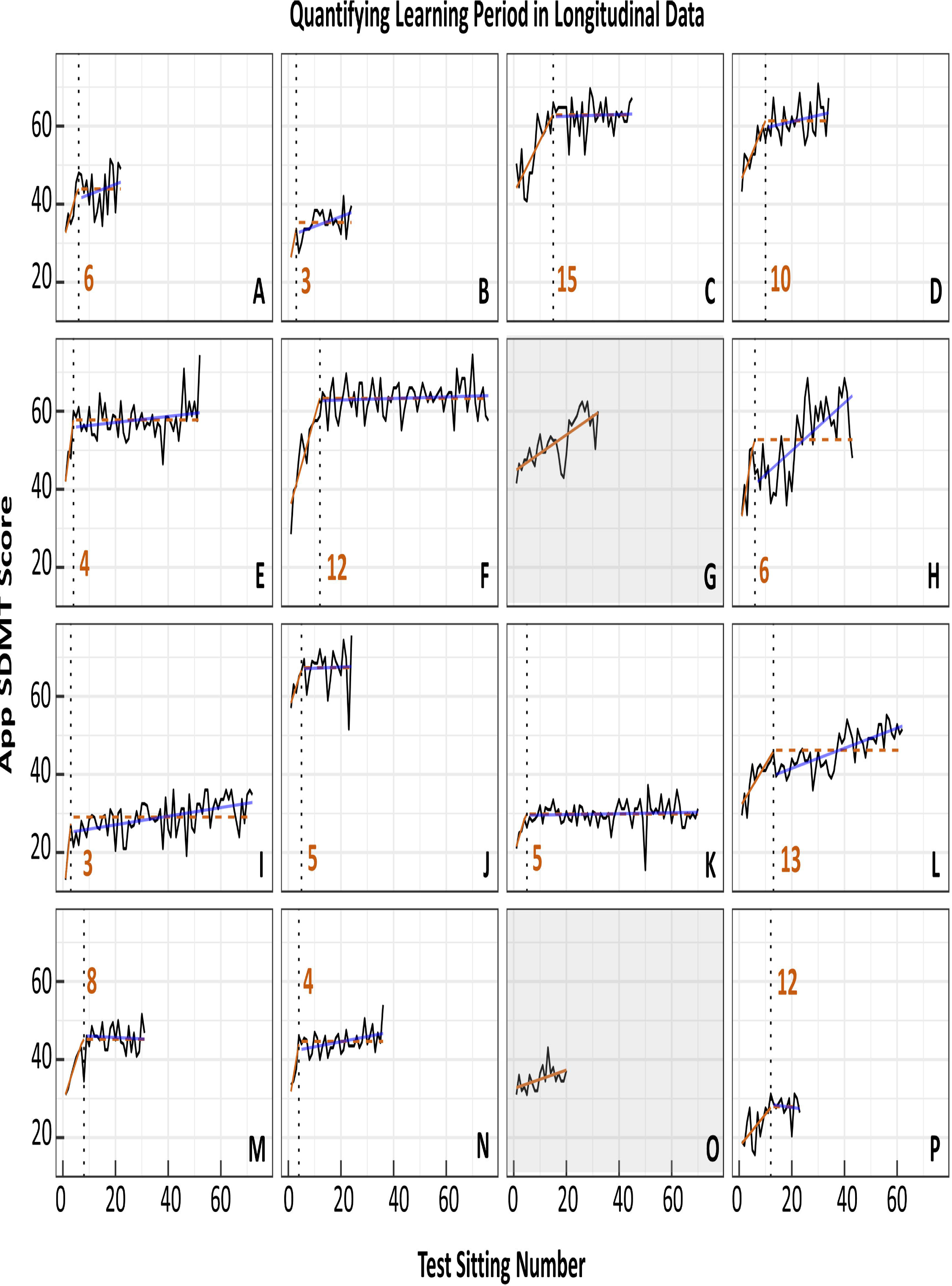
Nonlinear regression can be used to identify the inflection point in longitudinal data where learning stops occurring. Out of 16 participants with longitudinal data (≥ 20 test sittings), nonlinear regression was able to identify the inflection point in 14 individuals. The solid orange line indicates the period where the individual is still learning. The dotted black line signifies the point learning inflection point that the algorithm identified. The dashed orange line denotes the algorithm’s assumption of the data’s regression line after the learning period. The solid blue line represents the actual regression line of the data after the learning period. On average, learning stops occurring after 8 sittings.

Out of 16 individuals (1 HV, 15 MS) who have completed at least 20 test sittings, non-linear regression identified practice effects in 14 individuals. The non-linear regression failed to identify infliction point in the remaining two subjects (Fig. 8, panels G and O). It is plausible that these individuals are still experiencing learning effect (due to positive slope of linear regression) that did not peak even after 20 test repetitions.

In individuals whose practice effects were quantifiable, the effects stopped after 8 test sittings on average (ranging from 3 to 15 sittings). Spearman correlation coefficients for post-practice effect longitudinal data showed that 3 out of 16 had strong evidence of a continuous (albeit much slower) improvement of SDMT test scores over time (individuals H, I, L; p < 0.0001).

#### Smartphone SDMT scores have good intra-individual reliability

A mixed model intraclass correlation coefficient (ICC) analysis showed that in all the longitudinal data, including the period where practice effect was present (learning period), 87% of the total variance in the smartphone SDMT score was accounted for by within-patient data clustering (ICC = 0.87, Fig. 9A). When the learning period was removed, the ICC value increased to 0.90 (Fig. 9B), indicating very good reproducibility of within-subject results in comparison to between-subjects variance.

**Figure 9.**
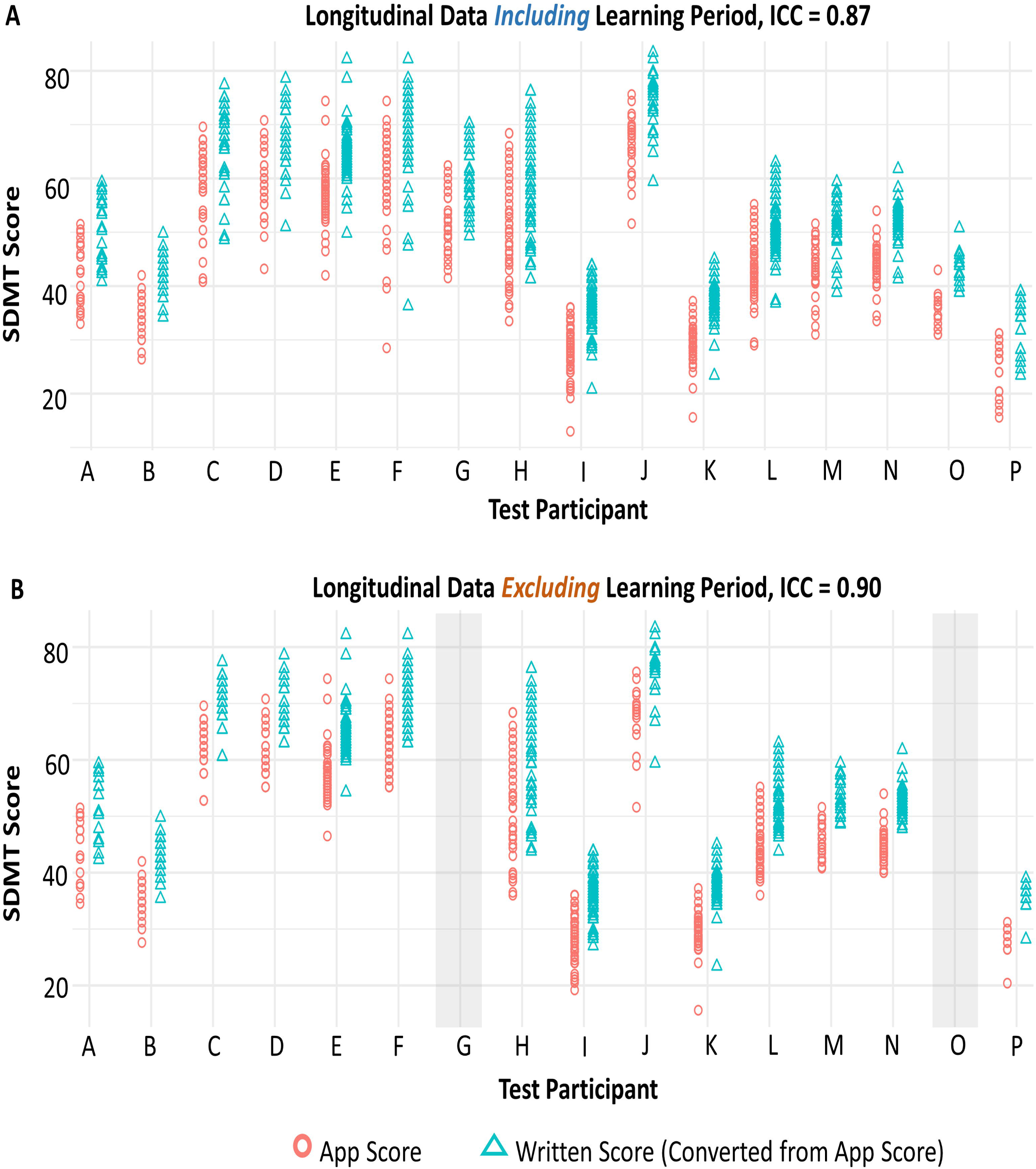
Longitudinal data exhibits good reliability. **(A)** With the learning period included, all longitudinal data has an intraclass correlation coefficient (ICC) of 0.87, which means that data for the same individual tend to cluster highly with each other and therefore, the data has good intraindividual reliability. **(B)** Without the learning period included, the ICC value increases to 0.90. In both panel A and B, the written SDMT scores (converted by adding 7.8 points to all app SDMT scores) have differences that span much larger than the current clinically significant change threshold of 4 points.

#### Longitudinal smartphone SDMT results enable a data-driven approach for defining clinically meaningful decline in cognitive processing speed

Already in the introduction we have alluded to the fact that currently, a 4-point decline in SDMT is considered “clinically meaningful” (5-7). However, when testing test-retest reliability by performing two trials of app based SDMT in one sitting, we observed intra-individual variance much greater than 4 points (Fig. 3A). Likewise, in MS subjects who perform SDMT weekly at home and whose linear models demonstrated either stable or slowly (but significantly) improving performance even after eliminating initial data affected by learning effect (Fig. 8), we observed intra-individual variability greatly exceeding 4 SDMT points (Fig. 9). None of these longitudinal testers reported MS relapse or significant deterioration of their neurological functions.

A plausible hypothesis to explain the discrepancy between the accepted 4-point SDMT threshold for clinically meaningful decline in traditional SDMT and our observations that the test-retest variance in app-SDMT greatly exceeds this threshold is that app based SDMT may be inherently noisier than traditional written SDMT.

We formally evaluated this hypothesis, even though it was not supported by the data presented thus far (i.e., app based SDMT had stronger, reproducible correlation with clinician-derived disability scores or relevant MRI features than written SDMT). First, we plotted the intra-individual differences between two written SDMT tests collected less than 190 days apart in 112 available MS patients (Fig. 10A, Table 2) and observed Gaussian distribution of these differences with mean of −0.7 and two standard deviations of 10.3. This variance is similar to what we observed in the test-retest Bland-Altman (Fig. 3A) of weekly longitudinal app SDMT sampling.

**Figure 10.**
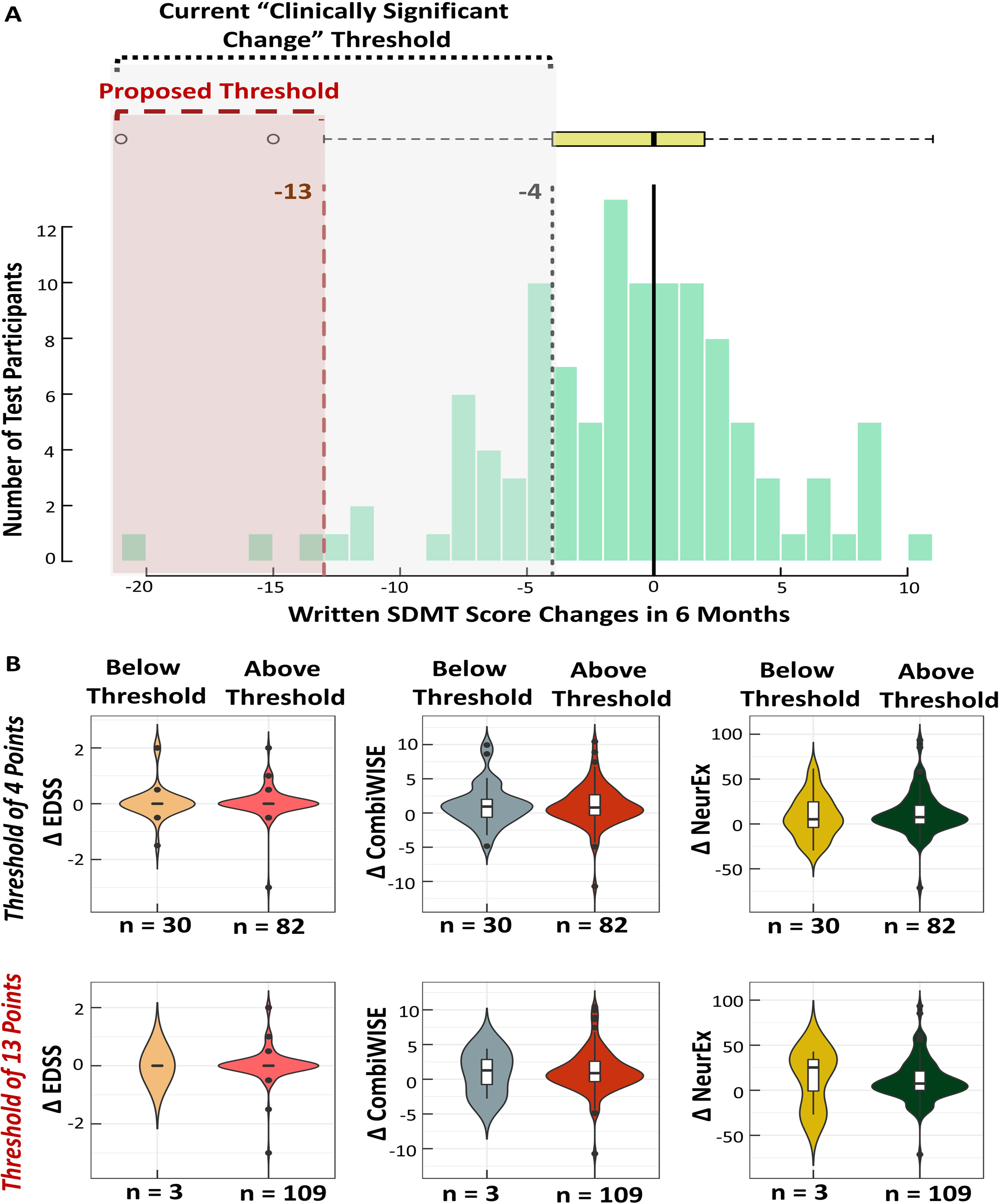
Proposing a new clinically significant change threshold for the written SDMT. **(A)** Distribution of written score differences for participants who completed the written SDMT in the course of 6 months. Using the current clinically significant score decrease of 4 points, approximately 24% of participants would have been falsely classified as having meaningful decline in their cognitive processing speed. We propose that this threshold should be increased to 13 points, or (average difference) – (1.5 × IQR of the differences). **(B)** Comparison of changes in disability scores for individuals who are above or below the defined clinically significant threshold. At the 4-points threshold, there is no significant difference in disability score changes between individuals who are below and above the threshold. At the 13-points threshold, while there is also no significant difference in disability changes between individuals who fall above or below the threshold, the disability changes are much larger in CombiWISE and NeurEx. The lack of statistical significance may be due to the low number of individuals who fall below the 13-points threshold.

Nevertheless, we formally tested if the 4-point written SDMT decline identified MS patients who progressed on disability scales capable of identifying MS progression in intervals as short as 6 months, such as the Combinatorial Weight-Adjusted Disability Scale (CombiWISE; continuous scale ranging from 0 to 100) (37) and NeurEx (Continuous scale ranging from 0 to theoretical maximum of 1349)(35). We also analyzed EDSS data, although the mean/median EDSS change over 6 months was expectedly zero (Fig. 10B). In contrast, both CombiWISE and NeurEx demonstrated measurable disease progression in the same group of patients (Δ CombiWISE +0.9 versus +0.8; Δ NeurEx + 5.2 versus +7.5).

Thirty MS patients (26.8%) fulfilled 4-point SDMT decline criterion, but we observed no statistically significant differences in either CombiWISE or NeurEx changes between MS patients who fell below and above this threshold (Fig. 10B; Δ CombiWISE comparison: p = 1.0; Δ NeurEx comparison: p = 0.4). When we chose a new threshold based on the distribution of differences in 6 months SDMT values (Average – 1.5 × Interquartile Range (IQR) = 13-point decline) we identified only 3 MS subjects (2.7%) who fulfilled this threshold, which uses statistically-accepted definitions of change that exceeds test-retest variance. This cohort is too small to obtain reliable statistical results, but we did observe that median CombiWISE and NeurEx changes were higher in this small group in comparison to the remaining 109 subjects with MS (Δ CombiWISE +1.3 versus +0.9; Δ NeurEx + 25.3 versus +7.5).

These results do not support the tested hypothesis that app based SDMT may be inherently “noisier” than traditional written SDMT. Instead we conclude that both SDMT tests have very similar variance, requiring difference of 13 to 14 SDMT points to accurately identify decline that exceeds test-retest fluctuation in SDMT performance when comparing only two SDMT tests within the same subject.

Next, we hypothesized that more frequent (granular) collection of data, permitted by the smartphone app may lower this threshold of meaningful deterioration when comparing period averages. The Gaussian distributions of test-retest data (for app based SDMT) or 6-month SDMT changes for the written test suggests random distribution of performance noise, that could be limited by averaging multiple weekly tests within individual patients, akin to “repeated measures”. To test this hypothesis, we compared, within our n = 14 post-learning longitudinal cohort, the variance of the app SDMT test differences when we compared single adjacent tests, with average of 2, 3 or 4 adjacent tests (Fig. 11). We observed expected decline in variance, from 14.4 SDMT points for single test comparisons to 9.5 points for average of 2 adjacent tests, 7.7 points for average of 3 adjacent tests and finally 6.2 points for average of 4 adjacent tests. We conclude that granular collection of SDMT data offers advantage over episodic collection of SDMT data in the clinic, as averaging multiple test repetitions lowers the threshold for identifying true decline in test performance.

**Figure 11.**
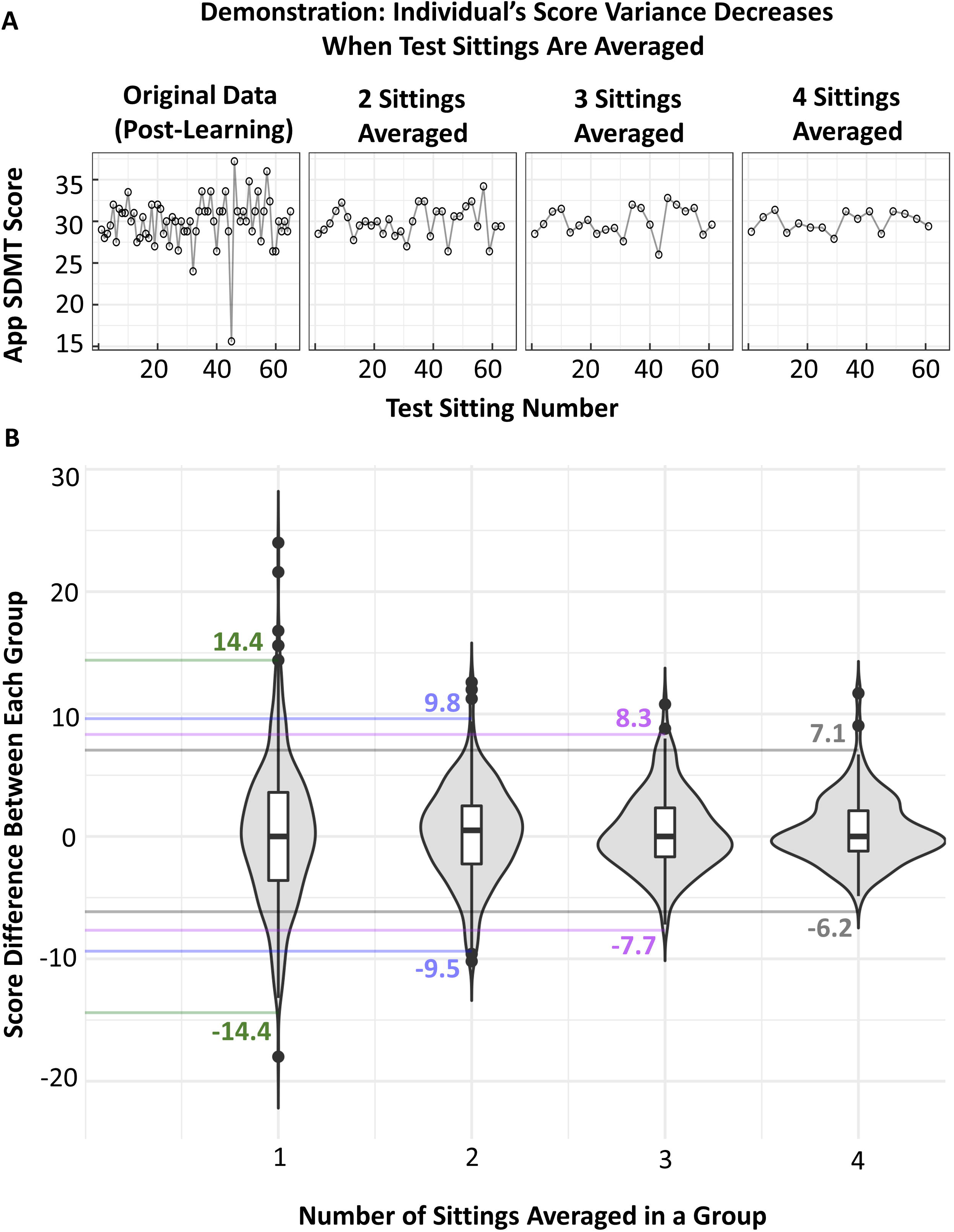
Determining the clinically significant change threshold for the app based SDMT. **(A)** A demonstration of how the significant thresholds are calculated in the app SDMT. For each individual, the score differences between one sitting, two sittings averaged, three sittings averaged, and four sittings averaged were calculated. **(B)** Plotting these differences allows for identification of the clinically significant change thresholds that should be used when the participant’s score changes are based on single test sittings or multiple sittings averaged together.

## Discussion

Neurological examination is a powerful tool to correctly identify, localize, and grade neurological deficits. However, socio-economic pressures are limiting access of neurological patients to this essential tool. The supply/demand for fully trained neurologists is critically low in many developing countries and is predicted to approach critical shortage in the USA this decade (1). The existing payer system forces neurologist to see follow-up patients in 10-15 minutes, while detailed performance of neurological examination requires 40-60 minutes. Consequently, most neurological patients are deprived of the benefit to monitor their disease using historically proven methodology.

Because it is unlikely that this situation will improve, it is imperative to find alternative ways to obtain reliable measurements of neurological disability. The “digital health” revolution spurred development of many “medical apps” adapted for smartphones or tablets. Some, like ours, are intended to address the aforementioned void. However, a recent conference on digital heath (38) concluded that technical development of these apps is easy in comparison to validating their value for intended use, in accordance with FDA guidance: “General principles of software validation; Final guidance for industry and FDA staff” (39). Consequently, most of these medical apps are released to the public without either formal software verification (i.e., consistency, completeness and correctness of the software) and validation (i.e., providing objective evidence that software specifications conform to user needs and intended uses). This reality may, paradoxically, stifle needed innovation, as release of multiple apps targeting the same disease fractionates affected population of patients and, if not delivering promised value, may eventually even antagonize them against the use of digital technologies.

As NIH investigators, our development and validation efforts are publicly funded and the resulting products are released free of charge to non-profit entities, and, after validation of the entire MS Test Suite, to the public. In accordance with aforementioned FDA guidance, our goal is to deliver an integrated and fully validated tool that is not only optimized from the standpoint of software verification, but also provides data-driven optimization of the outcome(s) derived from each test contained in the Suite, validation against gold-standard of neurological examination, and objective measurement of CNS tissue destruction, as is presented here for the SDMT app. Although not affecting scientific value of presented results, we do recognize that public funding and resultant free distribution of the validated tool may in fact limit its broad utility, as our research group will not be able to provide long-term maintenance of the software and needed customer support to patients and neurologists. Hopefully, private-public partnership(s) may solve this issue.

We focused our developmental efforts on smartphones instead of tablets due to the broad availability of the former, including in developing countries, where the shortage of neurologists is especially pressing. Adaptation of existing tests, like the SDMT, often developed and used in paper formats (letter or A4) to much smaller smartphone screens requires greater involvement of visual system/eye-hand coordination and hand dexterity from the tester, as revealed by our side-by-side comparison of the traditional written SDMT and its smartphone adaptation. This is a shortcoming in comparison to tablet-based apps that offer much larger screens. For example, we acknowledge that our smartphone-based SDMT adaptation achieved lower agreement (measured by Bland-Altman plot) with traditional SDMT than published, tablet-based, oral SDMT adaptation in the Cognitive Assessment for Multiple Sclerosis iCAMS (40). It is unclear if this stronger agreement would also be observed if iCAMS SDMT was administered in a patient-autonomous manner, without the involvement of a trained administrator, which is the ultimate goal of our MS Test Suite.

Nevertheless, as illustrated in this article, if the complete neurological examination (e.g., digitalized in NeurEx) is matched to the app test, it is possible to correctly map contributions of these diverse neurological functions to the given test, measure performance of contributing neurological functions in other apps of the MS Test Suite, and mathematically adjust for measured disabilities as covariates to isolate the tested neurological domain. This then leads to stronger correlation of optimized app-derived outcome(s) with the targeted neurological function or with objective measures of CNS tissue destruction that represents anatomical substrate of measured disability in comparison to the analogue version of the same test (Fig. 6). Using this methodology, the app SDMT results, when adjusted for visual disability and DH dexterity, explained a stunning 74% of the variance in the EN linear regression model against a composite of brain atrophy and T2LL in the independent validation cohort. This result is quite remarkable and validates the ability of smartphone SDMT to reliably measure MS-related brain injury.

Importantly, digitalization of the test offers advances not available for traditional tests, such as randomization of the symbol-digit code, algorithmic identification of the learning effect, granular longitudinal sampling permissive to averaging temporary-adjacent results for minimizing performance noise, thus providing accurate “baseline” for sensitive identification of the future true (i.e., disease-related) decline in test performance.

Indeed, our use of non-linear regressions on the level of individual subjects yielded identical result (i.e., stabilization of the learning effect after 8 trials) that was previously measured on a group level in natalizumab-treated MS patients (41). Identification of the peak of learning effect in individual subjects will likely lead to more accurate data, especially when the MS Test Suite is applied in clinical practice rather than in drug development.

Unexpectedly, we observed that test-retest within-subject variance in both app based and traditional SDMT greatly exceeded previously suggested threshold of 4 SDMT points indicative of “clinically meaningful change”. Unfortunately, papers that proposed 4 SDMT points threshold did not measure test-retest variance (5-7). When using this proposed threshold, 24% of MS patients in our cohort would be labeled as having “clinically-meaningful progression on SDMT test” over a short 6-month interval, which is unrealistic. Applying the same logic, even greater proportion of MS patients would have clinically meaningful improvements of cognitive functions over identical short interval. Indeed, we did not observe any differences on highly sensitive composite clinical scores (CombiWISE and NeurEx) between patients who fell below this threshold and those who did not over the course of 6 months.

Instead, our results show that a decline of 13-14 points on a single SDMT test is necessary to differentiate true cognitive decline from the noise in test performance. This data-driven threshold identified progression only in 2.7% of patients over identical 6-month interval and these patients also progressed on global measures of neurological disability. The 13-14 point threshold is quite insensitive but can be significantly decreased in granular sampling by averaging multiple temporally-related test results, which, akin to repeated measures, limits performance noise and leads to more reliable estimate of subject’s reaction time in comparison to the single measurement. For example, averaging 4 temporally-adjacent app SDMT results decreases the limit of clinically-meaningful decline to 6 SDMT points.

In conclusion, smartphone adaptation of the SDMT test allows for accurate measurement of reaction time in a patient-autonomous manner. Adjusting app SDMT results for relevant clinical covariates – some of which can be reliably measured by another smartphone test – leads to optimized outcome that correlates strongly with MS-related brain injury, measured by volumetric MRI.

## Data Availability

Data and script used for all analyses can be found in the github link (see supplementary file for the link).

https://github.com/vanessatmorgan/Bielekova-Lab-Code

## Data availability

Data used for all analyses can be found in the Supplementary Files.

## Ethics statement

This study was approved by and carried out in accordance with the recommendations of the Central Institutional Review Board of the National Institutes of Health (NIH). All subjects gave a written or digital informed consent in accordance with the Declaration of Helsinki.

## Author contribution

LP executed tests with patients in the clinic, performed all analyses detailed in the paper and wrote the first draft of this paper and all its figures. TH contributed to the initial app development, creation of app features, app updates, and maintenance of the app/cloud-based data storage. MV provided volumetric analyses of MRI data. PK calculated and exported clinical scores used in the correlation and elastic net analyses. BB developed the initial concept for the smartphone testing, guided/supervised all data analyses and app development, and edited the paper/figures for intellectual content.

## Acknowledgements

We would like to thank Dr. Atif Memon at the University of Maryland, College Park, for providing guidance in the initial development of the app. We would also like to thank Dr. Chris Barbour for suggesting and demonstrating how non-linear regression can be applied in our analyses. Finally, we would like to thank our clinical team, patients, and caregivers at the Neuroimmunological Diseases Section for partnering with us in this project. This study was funded by the Intramural Research Program of the National Institute Allergy and Infectious Diseases (NIAID) of the National Institutes of Health (NIH).

## References

1. Racette, B.A., et al., Supply and demand analysis of the current and future US neurology workforce. Neurology, 2014. 82(24): p. 2254–5.

2. Boukhvalova, A.K., et al., Smartphone Level Test Measures Disability in Several Neurological Domains for Patients With Multiple Sclerosis. Front Neurol, 2019. 10: p. 358.

3. Boukhvalova, A.K., et al., Identifying and Quantifying Neurological Disability via Smartphone. Frontiers in Neurology, 2018. 9.

4. Smith, A., Symbol Digit Modalities Test: Manual. 2002, Western Psychological Corporation: Los Angeles, California.

5. Benedict, R.H., et al., Characterizing cognitive function during relapse in multiple sclerosis. Mult Scler, 2014. 20(13): p. 1745–52.

6. Morrow, S.A., et al., Predicting loss of employment over three years in multiple sclerosis: clinically meaningful cognitive decline. Clin Neuropsychol, 2010. 24(7): p. 1131–45.

7. Pardini, M., et al., Isolated cognitive relapses in multiple sclerosis. J Neurol Neurosurg Psychiatry, 2014. 85(9): p. 1035–7.

8. Kosa, P., et al., NeurEx: digitalized neurological examination offers a novel high-resolution disability scale. Ann Clin Transl Neurol, 2018. 5(10): p. 1241–1249.

9. Sweeney, E.M., et al., OASIS is Automated Statistical Inference for Segmentation, with applications to multiple sclerosis lesion segmentation in MRI. Neuroimage Clin, 2013. 2: p. 402–13.

10. Parker, R.A., et al., Application of Mixed Effects Limits of Agreement in the Presence of Multiple Sources of Variability: Exemplar from the Comparison of Several Devices to Measure Respiratory Rate in COPD Patients. PLoS One, 2016. 11(12): p. e0168321.

11. R Development Core Team, R: A Language and Environment for Statistical Computing. 2018.

12. Sarkar, D., Lattice: Multivariate Data Visualization with R. 2008, New York: Springer.

13. Bates, D., et al., Fitting Linear Mixed-Effects Models Using [lme4]. Journal of Statistical Software, 2015. 67(1): p. 1–48.

14. Lüdecke, D., sjPlot: Data Visualization for Statistics in Social Science. 2020.

15. Kassambara, A., ggcorrplot: Visualization of a Correlation Matrix using ‘ggplot2’. 2019.

16. Signorell, A., et al., DescTools: Tools for Descriptive Statistics. 2019.

17. Wickham, H., Reshaping data with the reshape package. Journal of Statistical Software, 2007. 21(12).

18. Bakdash, J. and L. Marusich, rmcorr: Repeated Measures Correlation. 2018.

19. Urbanek, S. and J. Horner, Cairo: R Graphics Device using Cairo Graphics Library for Creating High-Quality Bitmap (PNG, JPEG, TIFF), Vector (PDF, SVG, PostScript) and Display (X11 and Win32) Output. 2019.

20. Simon, N., et al., Regularization Paths for Cox’s Proportional Hazards Model via Coordinate Descent. Journal of Statistical Software, 2011. 39(5): p. 1–13.

21. Kuhn, M., caret: Classification and Regression Training. 2020.

22. Pinheiro, J., et al., Linear and Nonlinear Mixed Effects Models. 2018.

23. Fox, J. and S. Weisberg, An [R] Companion to Applied Regression. Third ed. 2019, Thousand Oaks, CA: Sage.

24. Kassambara, A., ggpubr: ‘ggplot2’ Based Publication Ready Plots. 2019.

25. Wickham, H., ggplot2: Elegant Graphics for Data Analysis. 2016: Springer-Verlag New York.

26. Grolemund, G. and H. Wickham, Dates and Times Made Easy with [lubridate]. 2011. 40(3): p. 1–25.

27. Wickham, H. and L. Henry, tidyr: Tidy Messy Data. 2020.

28. Wickham, H., et al., dplyr: A Grammar of Data Manipulation. 2019.

29. Gandrud, C., DataCombine: Tools for Easily Combining and Cleaning Data Sets. 2016.

30. Wickham, H., J. Hester, and R. Francois, readr: Read Rectangular Text Data. 2018.

31. Ram, K. and H. Wickham, wesanderson: A Wes Anderson Palette Generator. 2018.

32. Lin, L.I., A concordance correlation coefficient to evaluate reproducibility. Biometrics, 1989. 45(1): p. 255–68.

33. Benedict, R.H., et al., Validity of the Symbol Digit Modalities Test as a cognition performance outcome measure for multiple sclerosis. Mult Scler, 2017. 23(5): p. 721–733.

34. Bland, J.M. and D.G. Altman, Statistical methods for assessing agreement between two methods of clinical measurement. Lancet, 1986. 1(8476): p. 307–10.

35. Kosa, P., et al., NeurEx: digitalized neurological examination offers a novel high-resolution disability scale. Annals of Clinical and Translational Neurology, 2018.

36. Rao, S.M., et al., Correlations between MRI and Information Processing Speed in MS: A Meta-Analysis. Mult Scler Int, 2014. 2014: p. 975803.

37. Kosa, P., et al., Development of a Sensitive Outcome for Economical Drug Screening for Progressive Multiple Sclerosis Treatment. Front Neurol, 2016. 7: p. 131.

38. in Digital Health: From Science to Application. 2019. Keystone, Colorado.

39. Administration, F.a.D., General Principles of Software Validation; Final Guidance for Industry and FDA Staff. 2002.

40. Beier, M., et al., iCAMS: Assessing the Reliability of a BICAMS Tablet Application. International Journal of MS Care Preprint, 2019.

41. Roar, M., Z. Illes, and T. Sejbaek, Practice effect in Symbol Digit Modalities Test in multiple sclerosis patients treated with natalizumab. Mult Scler Relat Disord, 2016. 10: p. 116–122.

